# Apolipoprotein E genotype, lifestyle and coronary artery disease: gene-environment interaction analyses in the UK Biobank population

**DOI:** 10.1101/2020.01.29.20019620

**Authors:** Maxime M Bos, Lina de Vries, Patrick CN Rensen, Ko Willems van Dijk, Gerard Jan Blauw, Diana van Heemst, Raymond Noordam

## Abstract

**Background:** Carriers of the *APOE ε4* genotype have an increased risk for developing coronary artery disease (CAD), but there is preliminary evidence that lifestyle factors interact with *APOE* genotype on CAD risk. Here, we assessed the interactions of physical activity, oily fish intake and polyunsaturated fatty acid (PUFA) intake with *APOE* genotype on risk of incident cardiovascular disease in a large population of middle-aged individuals.

**Methods and Results:** The present study was embedded in the UK Biobank population and comprised 344,092 European participants (mean age: 56.5 years, 45.7% men) without a history of CAD. Information regarding physical activity, oily fish intake and PUFA intake was collected through questionnaires, and information on incident CAD through linkage with hospital admission records. Analyses were performed using Cox proportional hazard models adjusted for age and sex. From these analyses, higher physical activity level and a higher intake of oily fish were associated with a lower incidence of CAD. These associations were similar across all APOE isoform groups (p-values for interaction > 0.05). A higher PUFA intake was only associated with a lower CAD risk in *APOE ε4* carriers (hazard ratio: 0.76, 95% confidence interval: 0.62,0.90), however, no statistically significant interaction was observed (p-value_interaction_ = 0.137).

**Conclusion:** While higher physical activity, fish intake and PUFA intake all decreased the risk of CAD, no evidence for interaction of these lifestyle factors with *APOE* genotype was observed in UK Biobank participants. Interventions intended to reduce cardiovascular risk might therefore be similarly effective across the APOE isoform carriers.

## Introduction

Despite the introduction of cholesterol-lowering medication and improved revascularization treatments, coronary artery disease (CAD) is still one of the most common causes of morbidity and mortality in the general population^1^. Much of the research in this area is focused on disentangling the pathophysiology of CAD and on the identification of novel targets for disease prevention. Large initiatives have been undertaken to investigate the genetics of CAD pathogenesis^2^. Genetic variation in the *APOE* gene has been widely recognized to increase the risk of CAD, which has also been confirmed by genome-wide association studies^2-4^. There is a standing notion that interactions between lifestyle and genetics may affect the response to cholesterol lowering medication and the susceptibility to CAD ^5^.

In order to explore the biological mechanisms via which lifestyle may modify disease traits, gene-environment interactions have been investigated^6-10^. We recently reviewed the current evidence on the existence of *APOE*-lifestyle interactions in the development of age-related diseases, including CAD, and argued that the beneficial effect of high physical activity and a high intake of oily fish might be largest for *APOE* ε4 carriers^11^. A population-based cross-sectional survey performed in 1,708 randomly selected participants aged 35 to 74 years, showed that a high intensity level of physical activity was associated with an increase of high-density lipoprotein (HDL) cholesterol and a decrease of triglyceride levels, specifically in *APOE* ε4 carriers^12^. Furthermore, It has been hypothesized that specifically PUFA intake may have a beneficial effect in *APOE* ε4 carriers. Small-scale intervention studies indeed indicate that polyunsaturated fatty acid (PUFA) intake and physical activity may have specific beneficial effects on CAD (risk factors) in *APOE* ε4 carriers^13-15^. However, a study done in 136,701 high CAD risk participants did not indicate that the *APOE-*lipid level association was modified by a high omega-3 intake^16^.

Given these contrasting results, there is a need for large scale studies examining effect modification of the *APOE* gene by lifestyle factors such as oily fish intake, PUFA and physical activity on incident CAD. Recently, large studies like the UK Biobank cohort that are well-powered and have data available on large numbers of incident CAD cases have become available as a platform to address such research questions In the present study, we assessed whether the association between *APOE* genotype and incident CAD risk is modified by oily fish and PUFA intake and physical activity levels in middle-aged individuals without a history of CAD from the UK Biobank cohort.

## Methods

### Study setting and population

The UK Biobank cohort is a prospective general population cohort. Baseline assessments took place between 2006 and 2010 in 22 different assessment centers across the United Kingdom ^17^. A total of 502,628 participants between the age of 40 and 70 years were recruited from the general population. Invitation letters were sent to eligible adults registered to the National Health Services (NHS) and living within a 25 miles distances from one of the study assessment centers. At the study assessment center, participants completed a questionnaire through touchscreen that included topics as sociodemographic characteristics, physical and mental health, lifestyle and habitual food intake. All participants from the UK Biobank cohort provided written informed consent, and the study was approved by the medical ethics committee. The project was completed under project number 32292.

In the present study, genotyped European-ancestry participants without a history of cardiovascular disease were followed till the development of the study outcome, death or the end of the study period (March 31, 2017; N = 363,745). Participants with missing data on questionnaire-based oily fish intake (N = 1,654) and physical activity frequency per week (N = 16,432) were excluded from the study. In total, the present study was conducted in 345,659 participants. Additionally, we performed analyses in a subsample of the study population with data on polyunsaturated fatty acids (PUFA) intake (N = 52,478).

### APOE genotyping

UK Biobank genotyping was conducted by Affymetrix using a bespoke BiLEVE Axium array for approximately 50,000 participants; the remaining participants were genotyped using the Affymetrix UK Biobank Axiom array. All genetic data were quality controlled centrally by UK Biobank resources. More information on the genotyping processes can be found online (https://www.ukbiobank.ac.uk). SNPs in *APOE* determining the isoform (notably rs7412 and rs429358) were directly genotyped. Both genetic variants were in Hardy-Weinberg equilibrium (p-value > 0.05). As a reference group, we used participants who are homozygous for the *APOE* ε3 allele (ε3/ε3 genotype). The *APOE* ε4 group consisted of individuals with the genotype ε3/ε4 and ε4/ε4. The *APOE* ε2 group consisted of individuals with the genotype ε2/ε3 and ε2/ε2. We excluded participants with other (rarer) genotypes (e.g. ε2/ε4).

### Lifestyle exposures

Via touchscreen questionnaires, information on the frequency of oily fish intake and physical activity per week was collected^18^. Via the same questionnaires, we determined the number of days per week at which participants had more than 10 minutes of vigorous physical activity^19^, which was defined as “doing physical activity that made you sweat or breathe hard”. Groups for oily fish intake and physical activity were divided based on whether individuals reported oily fish intake of at least once per week or were active for at least one day per week. In order to assess whether there is a dose-response relationship for oily fish intake and physical activity on a lower CAD incidence, we formed groups based on whether individuals had oily fish intake or physical activity only once, twice, or three or more times per week. As a reference group we used those individuals who reported to not have any intake of oily fish or were not active for any day of the week.

In a subset of the population, the frequency of intake of 200 consumed food items and drinks over the previous 24 hours was collected with a 24-hour dietary recall questionnaire (25) based on which the average intake of macro- and micronutrients was calculated. Information regarding polyunsaturated fatty acid (PUFA) intake was obtained via this questionnaire. The groups for PUFA intake were based on the median PUFA intake in which a higher than median PUFA intake was considered high and a lower than the median intake was considered as low and used as a reference group in our analyses.

### Cardiovascular disease outcomes

Information on incident cardiovascular disease was collected through information from the data provided by the NHS record systems. Diagnoses were coded according to the International Classification of Diseases (ICD)^17^. Here, the study outcome was CAD which we defined as: angina pectoris (I20), myocardial infarction (I21), acute and chronic ischemic heart disease (I24 and I25) and stroke (I63 and I64).

### Covariates

Body Mass Index (BMI) was calculated by dividing the weight in kilograms by the height in meters squared, which were measured objectively at the study center. Participants were asked to remove shoes and heavy outer clothing before weighting. During the visit of the assessment center, participants completed touchscreen questionnaires regarding smoking status (never, previous or current), frequency of alcohol consumption, disease status (e.g. diabetes mellitus) and medication usage (lipid lowering medication and blood pressure lowering medication).

### Statistical analyses

Characteristics of the study population were examined at baseline and expressed as means (standard deviations), medians (interquartile ranges; for non-normally distributed variables only), and proportions.

We examined the association between physical activity, oily fish intake and PUFA intake on incident CAD in a population without a history of CAD using cox proportional hazard models adjusted for age and sex in R (version 3.6.1) using the survival package (version 2.44-1.1)^20, 21^. Results were visualized using the R-based packages ggplot2, survminer and the metafor package^22-24^. Participants were followed till the first CAD event, death or the end of the study period (March 31, 2017), whichever came first. Results are presented as the hazard ratios with the accompanying 95% confidence intervals. Since individuals who are at high-risk for development of CAD may alter their lifestyle, in a sensitivity analysis we excluded participants who used lipid lowering medication, blood pressure lowering medication or had a clinical diagnosis of diabetes mellitus. In order to formally test for an interaction, we added an interaction term between the *APOE* genotype and the lifestyle factor to the model.

## Results

### Characteristics of the study population

A total of 345,659 white Caucasian participants were included in the present study. In **Table 1**, the population characteristics for the study population are presented stratified by APOE isoform. In general, study characteristics are comparable between subgroups. However, statin use, total cholesterol levels and LDL-cholesterol are lower in individuals with the *APOE* ε2 genotype as compared to *APOE* ε3 carriers and higher for *APOE* ε4 carriers. In **Supplementary Table 1**, the population characteristics are presented when stratified based on lifestyle factors. Individuals with a low fish intake are slightly younger than those with a high fish intake. Moreover, individuals with a high physical activity level are more often males, have a lower statin use, a lower use of blood pressure lowering medication and have less diabetes as compared to individuals with a low physical activity level.

**Table 1.** Baseline characteristics of the participants in the UK Biobank, stratified by *APOE* genotype

|  | **APOE ε2** | **APOE ε3** | **APOE ε4** |
| --- | --- | --- | --- |
|  | *N*= 45,570 | *N*= 207,909 | *N*= 92,180 |
| **Demographics** |  |  |  |
| Age in years, mean (SD) | 56.5 (8.0) | 56.4 (8.0) | 56.3 (8.0) |
| Sex, N (%male) | 20,445 (44.9) | 92,612 (44.5) | 40,930 (44.4) |
| **Lifestyle variables** |  |  |  |
| BMI, mean (SD) | 27.3 (4.7) | 27.2 (4.7) | 27.1 (4.7) |
| Smoking status, N (%current) | 4,199 (9.2) | 19,221 (9.2) | 8,236 (8.9) |
| Oily fish intake in days per week, median (IQR) | 2 [1-2] | 2 [1-2] | 2 [1-2] |
| 10 minutes of vigorous PA in days per week, median (IQR) | 1 [0-3] | 1 [0-3] | 1 [0-3] |
| **Cardiovascular risk factors** |  |  |  |
| Statin use, N (%yes) | 4,613 (10.1) | 29,120 (14.0) | 15,394 (16.7) |
| Blood pressure lowering medicines use, N (%yes) | 8,039 (17.6) | 37,763 (18.2) | 16,823 (18.3) |
| Diabetes, N (%yes) | 1,955 (4.3) | 8,613 (4.1) | 3,492 (3.8) |
| Cholesterol in mmol/L, mean (SD) | 5.4 (1.0) | 5.8 (1.1) | 5.9 (1.2) |
| HDL-cholesterol in mmol/L, mean (SD) | 1.5 (0.4) | 1.5 (0.4) | 1.4 (0.4) |
| LDL-cholesterol in mmol/L, median (IQR) | 3.2 [2.7 – 3.7] | 3.6 [3.0 – 4.2] | 3.7 [3.2 – 4.3] |
| Triglycerides in mmol/L, median (IQR) | 1.5 [1.1 – 2.3] | 1.4 [1.0 – 2.1] | 1.5 [1.1 – 2.2] |

Abbreviations: BMI, body mass index; IQR, interquartile range; N, number; PA, physical activity; SD, standard deviation.

### Incident cardiovascular disease per APOE isoform

A total of 12,806 participants had an incident event of coronary artery disease (CAD) during a median follow-up period of 8.11 years. **Figure 1** depicts the event-free survival probability per *APOE* isoform. The highest event-free survival probability was observed in ApoE ε2 carriers and the lowest probability in ApoE ε4 carriers with accompanying hazard ratios of 0.93 (95% confidence interval (CI) 0.88 – 0.98) and 1.10 (95%CI: 1.06 – 1.14).

**Figure 1.**
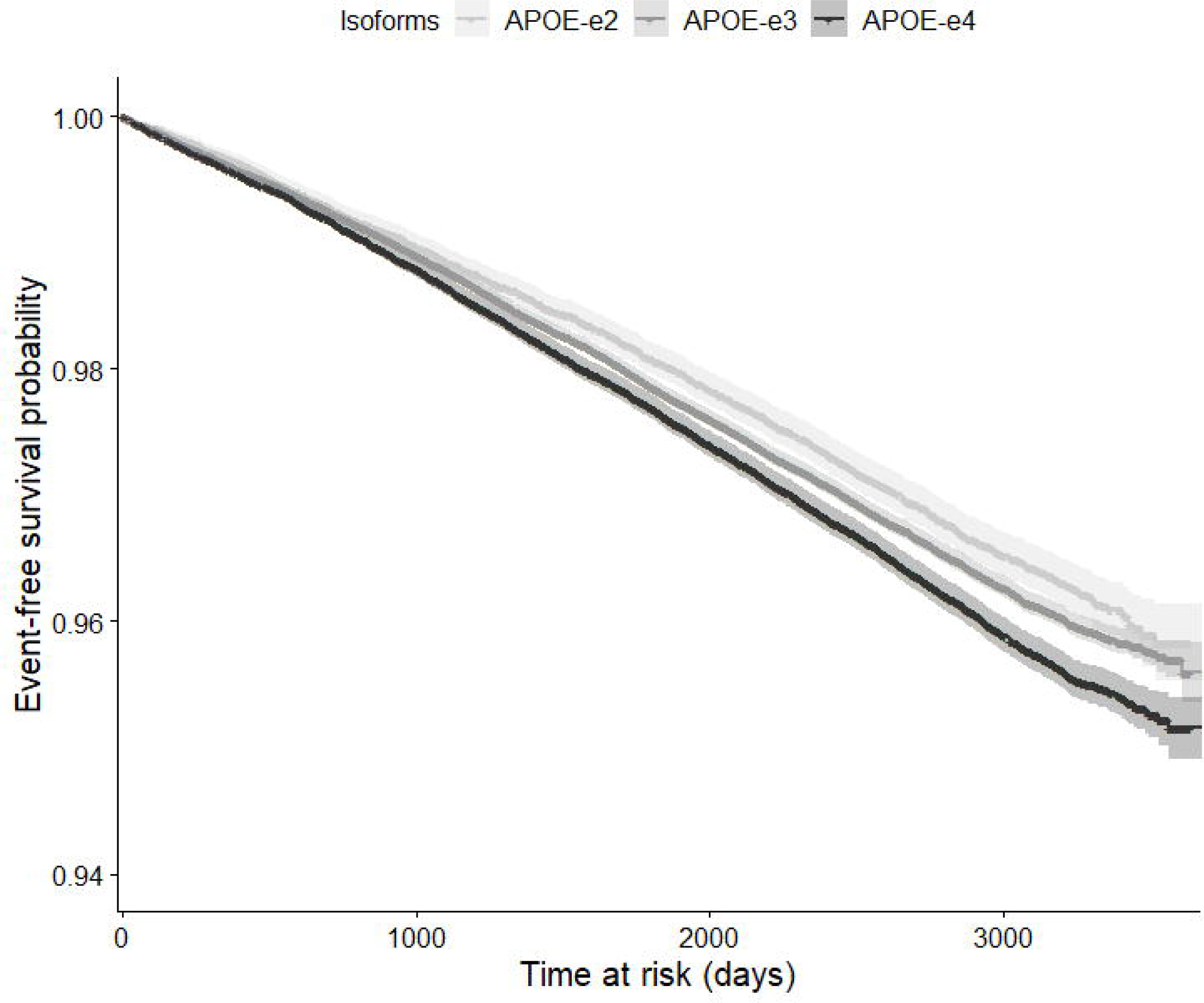
Cardiovascular disease-free survival per *APOE* isoform.

### Incident cardiovascular disease per lifestyle group

As shown in **Figure 2**, oily fish intake was associated with a lower incidence of CAD in carriers of the ε2 isoform (hazard ratio (HR): 0.78 [95% confidence interval (CI): 0.66 – 0.91]), in carriers of the ε3 isoform (HR: 0.76 [95%CI: 0.71 – 0.82]), and in carriers of the ε4 isoform (HR: 0.83 [95%CI: 0.74 – 0.92]). Physical activity was associated with a lower CAD incidence in *APOE* ε2 carriers (HR: 0.81 [95%CI: 0.73 – 0.89]), in *APOE* ε3 carriers (HR: 0.77 [95%CI: 0.73 – 0.80]) as well as in ApoE ε4 carriers (HR: 0.76 [95%CI: 0.71 – 0.81]). Here, we did not find evidence for an interaction with fish intake or physical activity with *APOE* genotype on the incidence of CAD. A high PUFA intake was associated with a lower incidence of CAD (HR: 0.76 [95%CI: 0.62 – 0.90], only in ε4 carriers, however no interaction was observed (**Figure 2**). Moreover, no clear dose-response relationship was observed for either fish intake or physical activity and CAD risk (**Supplementary Figure 1**). However, when the frequency of fish intake or physical activity is taken into account, an interaction for once a week of physical activity in the group with ApoE ε2 and ApoE ε3 and an interaction for once a week of fish intake in the group with ApoE ε3 and ApoE ε4 was observed (**Supplementary Figure 1**).

**Figure 2.**
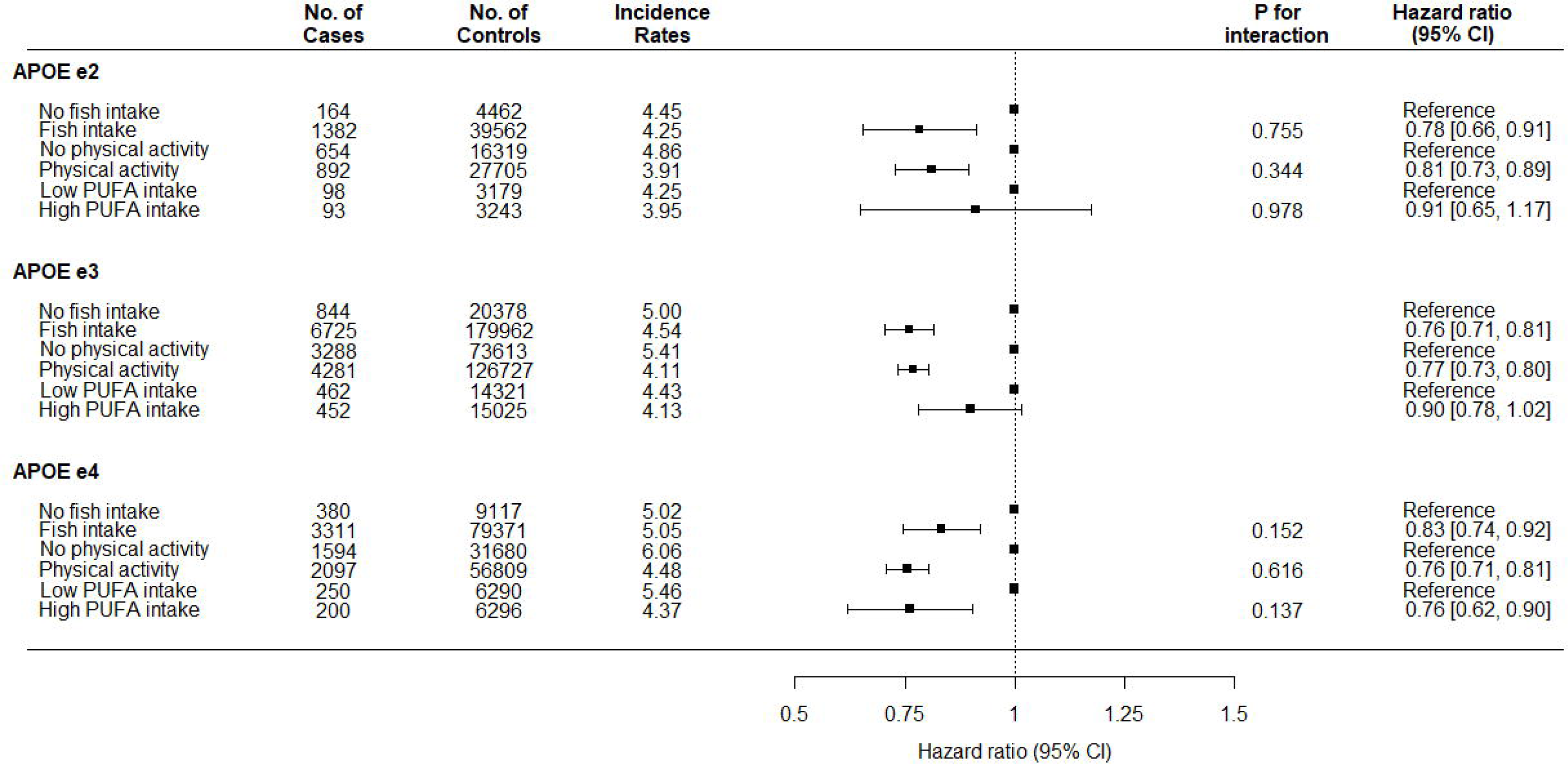
Hazard ratios for CAD incidence for fish intake, physical activity and polyunsaturated fatty acid (PUFA) intake, stratified by *APOE* isoform.

As a sensitivity analysis, we excluded participants who used lipid-lowering medication, blood pressure-lowering medication or with self-reported diabetes mellitus. Results were comparable to those obtained in our previous analyses (**Supplementary Figures 1 and 2**). Results were similar when we stratified by sex (results not shown).

## Discussion

In the present study, we assessed whether there is evidence for lifestyle-*APOE* interactions on incident CAD in middle-aged individuals of the UK Biobank. Here, we reported no evidence for interaction between fish intake, physical activity and polyunsaturated fatty acid (PUFA) intake with *APOE* genotype on incident CAD in the large European UK Biobank population. However, we showed that, independent of *APOE* genotype, a higher intake of fish and a higher physical activity level both associated with a lower CAD risk. A higher PUFA intake was only associated with a lower CAD risk in *APOE* ε4 carriers, however, no formal statistical interaction was observed.

ApoE ε4 has a different binding affinity for lipoprotein particles or the LDL-receptor than the other isoforms^25^. ApoE ε4 has a higher binding ability for triglyceride-rich lipoproteins, such as chylomicron remnants and very-low-density-lipoprotein (VLDL) particles, than for LDL particles^26^. This results in a diminished clearance of LDL-cholesterol, resulting in higher LDL-cholesterol levels as compared to carriers of APOE ε2 and ε3^27^. We hypothesized that carriers of the ε4 isoform may benefit differently from a higher physical activity and a higher intake of oily fish, however, we did not find evidence supporting this hypothesis. In the present study, for individuals with a high physical activity level, the incidence rate for CAD was lower than for those with a low physical activity level. These lower incidence rates were similar across the different *APOE* carrier groups. Therefore, a higher physical activity is likely to be beneficial irrespective of *APOE* genotype. Moreover, in our study the incidence rate of CAD decreased with a higher oily fish intake, in carriers of all ApoE isoforms and was not higher in carriers of the ε4 isoform. Interestingly, a higher intake of PUFA associated with a lower CAD incidence only in *APOE* ε4 carriers points towards a possible lifestyle-*APOE* interaction on incident CAD. One possible explanation is that intake of PUFA may result in a lower concentration of VLDL and thereby may increase the uptake of LDL by the liver, resulting in lower LDL levels^11^. However, these observations likely have no clear benefit at a clinical level. Since the group with information regarding PUFA intake was relatively small this may have resulted in limited statistical power and, therefore, further research to investigate the effect on a larger population level is warranted.

Although *APOE*-lifestyle interactions on cognitive function have been hypothesized as well^11^, no significant interaction with cognitive function was observed in the UK Biobank population previously^28^. The lack of a *APOE*-lifestyle interactions in this population may be a reflection of some sort of selection bias where the current study population includes in general more healthy *APOE* ε4 carriers. Indeed, there is evidence of a ‘healthy volunteer’ bias in the UK Biobank sample^29^. Moreover, a genome-wide association study on habitual physical activity in the UK Biobank identified *APOE* to be one of the strongest associations with physical activity^19^. The association was markedly stronger among older participants, therefore it may be that the older *APOE* risk allele carriers are particularly enriched for healthy lifestyle. One explanation could be that individuals with a known familial history of dementia and cardiovascular disease purposefully increase their physical activity levels and intake of oily fish. This bias may have resulted in an underestimation of the effect of exercise and fish intake. Alternatively, participants of the UK Biobank are still in good health or are slightly too young to show significant effects of the *APOE* genotype on CAD incidence. Moreover, additional lifestyle interactions, not covered in the present study, with *APOE* isoform may be possible. However, it should be noted that recent large-scale genome-wide interaction efforts were not able to identify interactions between physical activity, sleep duration, alcohol intake or smoking on blood lipid levels, limiting the potential of APOE-lifestyle interactions in the pathogenesis of CAD^6-10^.

A strength of this study is that we used data from the UK Biobank and therefore, to the best of our knowledge, this is the largest study to test for an interaction of lifestyle factors and *APOE* on incident CAD. However, the current analyses have only been performed in participants of Caucasian descent, thereby hampering the translation to individuals with a different ancestry. This is of particular importance since variability exists in *APOE*-related disease prevalence in different ancestry groups.

Based on our results, it is unlikely that there is a significant interaction between genetic variation in *APOE* and various lifestyle factors on the development of incident CAD in a population of middle-aged and older individuals free of CAD history. Therefore, it seems likely that interventions intended to reduce cardiovascular risk via increased physical activity and increased intake of oily fish will be equally effective in carriers of the APOE ε4 genotype as in the other *APOE* genotype carriers and may thus have considerable health benefits irrespective of *APOE* isoform.

## Data Availability

Data of the UK Biobank is available upon acceptance of a research proposal.

https://www.ukbiobank.ac.uk/

## Acknowledgements

The present UK Biobank project was conducted under project number 32292.

## Figure legends

**Supplementary Figure 1**. Hazard ratios for CAD incidence for categorized fish intake and physical activity, stratified by *APOE* isoform.

**Supplementary Figure 2**. Hazard ratios for CAD incidence for fish intake, physical activity and PUFA intake, stratified by *APOE* isoform with exclusion of individuals that used lipid-lowering medication, blood pressure-lowering medication or had a clinical diagnosis of diabetes mellitus.

**Supplementary Figure 3**. Hazard ratios for CAD incidence for categorized fish intake and physical activity, stratified by *APOE* isoform with exclusion of individuals that used lipid-lowering medication, blood pressure-lowering medication or had a clinical diagnosis of diabetes mellitus.

